# Optimizing Individual Targeting of Fronto-Amygdala Network with Transcranial Magnetic Stimulation (TMS): Biophysical, Physiological and Behavioral Variations in People with Methamphetamine Use Disorder

**DOI:** 10.1101/2023.04.02.23288047

**Authors:** Ghazaleh Soleimani, Christine A. Conelea, Rayus Kuplicki, Alexander Opitz, Kelvin O Lim, Martin P. Paulus, Hamed Ekhtiari

**Author notes:** **Corresponding Author:** Hamed Ekhtiari, MD, PhD, Laureate Institute for Brain Research, 6655 South Yale Ave. Tulsa, OK 74136.

## Abstract

**Background:** Previous studies in people with substance use disorders (SUDs) have implicated both the frontopolar cortex and amygdala in drug cue reactivity and craving, and amygdala-frontopolar coupling is considered a marker of early relapse risk. Accumulating data highlight that the frontopolar cortex can be considered a promising therapeutic target for transcranial magnetic stimulation (TMS) in SUDs. However, one-size-fits-all approaches to TMS targets resulted in substantial variation in both physiological and behavioral outcomes. Individualized TMS approaches to target cortico-subcortical circuits like amygdala-frontopolar have not yet been investigated in SUDs.

**Objective:** Here, we (1) defined individualized TMS target location based on functional connectivity of the amygdala-frontopolar circuit while people were exposed to drug-related cues, (2) optimized coil orientation based on maximizing electric field (EF) perpendicular to the individualized target, and (3) harmonized EF strength in targeted brain regions across a population.

**Method:** MRI data including structural, resting-state, and task-based fMRI data were collected from 60 participants with methamphetamine use disorders (MUDs). Craving scores based on a visual analog scale were collected immediately before and after the MRI session. We analyzed inter-subject variability in the location of TMS targets based on the maximum task-based connectivity between the left medial amygdala (with the highest functional activity among subcortical areas during drug cue exposure) and frontopolar cortex using psychophysiological interaction (PPI) analysis. Computational head models were generated for all participants and EF simulations were calculated for fixed vs. optimized coil location (Fp1/Fp2 vs. individualized maximal PPI location), orientation (AF7/AF8 vs. orientation optimization algorithm), and stimulation intensity (constant vs. adjusted intensity across the population).

**Results:** Left medial amygdala with the highest (mean ± SD: 0.31±0.29) functional activity during drug cue exposure was selected as the subcortical seed region. Amygdala-to-whole brain PPI analysis showed a significant cluster in the prefrontal cortex (cluster size: 2462 voxels, cluster peak in MNI space: [25 39 35]) that confirms cortico-subcortical connections. The location of the voxel with the most positive amygdala-frontopolar PPI connectivity in each participant was considered as the individualized TMS target (mean ± SD of the MNI coordinates: [12.6 64.23 -0.8] ± [13.64 3.50 11.01]). Individual amygdala-frontopolar PPI connectivity in each participant showed a significant correlation with VAS scores after cue exposure (*R*=0.27, *p*=0.03). Averaged EF strength in a sphere with r = 5mm around the individualized target location was significantly higher in the optimized (mean ± SD: 0.99 ± 0.21) compared to the fixed approach (Fp1: 0.56 ± 0.22, Fp2: 0.78 ± 0.25) with large effect sizes (Fp1: *p* = 1.1e-13, Hedges’g = 1.5, Fp2: *p* = 1.7e-5, Hedges’g = 1.26). Adjustment factor to have identical 1 V/m EF strength in a 5mm sphere around the individualized targets ranged from 0.72 to 2.3 (mean ± SD: 1.07 ± 0.29).

**Conclusion:** Our results show that optimizing coil orientation and stimulation intensity based on individualized TMS targets led to stronger electric fields in the targeted brain regions compared to a one-size-fits-all approach. These findings provide valuable insights for refining TMS therapy for SUDs by optimizing the modulation of cortico-subcortical circuits.

**Short Abstract:** *Background:* Prior research on drug addiction has linked the frontopolar cortex and amygdala coupling to drug cue reactivity/craving. However, one-size-fits-all approaches for transcranial magnetic stimulation (TMS) over frontopolar-amygdala have led to inconsistent results.

*Objective:* Here, we (1) defined individualized TMS target location based on functional connectivity of the amygdala-frontopolar circuit while people were exposed to drug-related cues, (2) optimized coil orientation for maximum electric field (EF) perpendicular to the individualized target, and (3) harmonized EF strength in targeted brain regions across a population.

*Method:* MRI data were collected from 60 participants with methamphetamine use disorders (MUDs). and examined the variability in TMS target location based on task-based connectivity between the frontopolar cortex and amygdala. using psychophysiological interaction (PPI) analysis. EF simulations were calculated for fixed vs. optimized coil location (Fp1/Fp2 vs. individualized maximal PPI), orientation (AF7/AF8 vs. optimization algorithm), and stimulation intensity (constant vs. adjusted intensity across the population).

*Results:* Left medial amygdala with the highest (0.31±0.29) fMRI drug cue reactivity was selected as the subcortical seed region. The location of the voxel with the most positive amygdala-frontopolar PPI connectivity in each participant was considered as the individualized TMS target (MNI coordinates: [12.6,64.23,-0.8]±[13.64,3.50,11.01]). Individualized frontopolar-amygdala connectivity showed a significant correlation with VAS craving scores after cue exposure (*R*=0.27, *p*=0.03). Averaged EF strength in a sphere with r=5mm around the individualized target location was significantly higher in the optimized (0.99±0.21V/m) compared to the fixed approach (Fp1:0.56±0.22V/m, Fp2:0.78±0.25V/m) with large effect sizes (Fp1*:p*=1.1e-13,Hedges’g=1.5, Fp2:*p*=1.7e-5,Hedges’g=1.26). Adjustment factor to have identical 1V/m EF strength in a 5mm sphere around the individualized targets ranged from 0.72-to-2.3 (1.07±0.29).

*Conclusion:* Our results show that optimizing coil orientation and stimulation intensity based on individualized TMS targets led to stronger harmonized electric fields in the targeted brain regions compared to a one-size-fits-all method that hopefully helps to refine future TMS therapy for MUDs.

## 1. Introduction

Substance use disorders (SUDs) are characterized by recurrent, compulsive, and excessive use of drugs that activate the brain’s reward system [1]. Previous work has implicated both the frontopolar cortex (medial prefrontal cortex, Brodmann Area 10) and the amygdala in substance-associated plasticity through the prefrontal-limbic system [2]. The amygdala is critical in responses to affective stimuli, and the frontopolar cortex is thought to be involved in tasks that require control of cognition and action [3]. Amygdala-frontopolar coupling is also recognized as a marker of early relapse risk in people with SUDs[4]. Both anatomical (white matter tracts) [5], [6], [7] and functional [8], [9], [10] connections between the frontopolar cortex and amygdala were reported in both human and animal studies. The functional connection between these two regions is dynamic and can be modulated with real-time fMRI amygdala neurofeedback [11], [12] or frontopolar transcranial magnetic stimulation (TMS) [13], [14].

The frontopolar cortex is considered a transdiagnostically relevant TMS target that induces local activation of neurons as well as transsynaptic activation of distal functionally/anatomically connected brain regions [15]. Previous TMS studies showed that excitatory/inhibitory stimulation of the frontopolar area can modulate cortico-subcortical circuits (e.g., frontopolar-amygdala) involved in drug cue reactivity and ultimately decrease drug-related behaviors such as drug craving and consumption [16]–[27]. For instance, it has been reported that inhibitory rTMS over the left frontopolar cortex can potentially revert hyperactivity in brain regions related to SUDs including the striatum, insula, and amygdala [19], [26], [28]. Although frontopolar TMS has already been applied clinically for SUDs, little is known about protocol optimization, especially as relating to the inter-individual variability [29], [30].

The integration of TMS with neuroimaging data, such as fMRI, has advanced our understanding of the functional correlates of TMS by revealing alterations in brain activity/connectivity patterns. This approach helps to utilize between-subject variations in response to stimulation for more precise positioning of the TMS coil over a therapeutic target [31], [32]. Previous TMS studies in SUDs used standard scalp-based targeting methods (e.g., EEG standard system) or neuronavigation systems which ignore the heterogeneity of the employed TMS dosage and functional brain network topography [15]. Current clinical practice for depression identifies the optimal stimulation target using TMS-fMRI integration, shifting the focus from brain anatomical regions to brain circuits at the individual level [32]–[34]. It has been observed that anti-depressant outcomes were better when TMS was applied at sites of the DLPFC that showed a stronger negative correlation with the sub-genual cingulate cortex. These findings were corroborated in several studies [35]–[37], and the individualized method augmented with depth-corrected stimulation intensity in an accelerated protocol recently received FDA clearance [38]. Similarly, investigating the effects of functionally guided, connectivity-based rTMS on amygdala activation were also investigated by individually targeting the prefrontal cortex in healthy young adults offered promising preliminary evidence that fMRI-informed targeting may provide a useful approach to treat network dysregulation [39], [40]. To improve stimulation accuracy even further, integrating functional connectivity maps with electric field (EF) simulation was also suggested by estimating the changes in connection strength based on inter-individual variability in the head and brain anatomy [30], [41]. However, less attention has been paid to optimizing stimulation methods in TMS for SUDs.

Inspired by previous functional connectivity maps integrated with EFs for TMS targeting, we believe that individualized stimulation approaches can precisely identify a coordinate in the frontopolar area to effectively modulate the amygdala-frontopolar circuit in individuals with SUDs. We applied this approach to a group of participants with methamphetamine use disorders (MUDs). We aimed to: (1) individualize TMS target location based on functional connectivity of the amygdala-frontopolar circuit during exposure to drug-related cues, (2) optimize coil orientation based on maximizing electric field (EF) perpendicular to the individualized target, and (3) harmonize EF strength in targeted brain regions across 60 participants with MUDs. Our computationally informed method seeks to optimize TMS protocols for SUDs via refined, individually targeted modulation of cortico-subcortical circuits.

## 2. Method

### 2.1. Participants

60 participants (all-male, mean age ± standard deviation (SD) = 35.86 ± 8.47 years ranging from 20 to 55) with MUD were recruited during the early abstinence phase (abstinence from methamphetamine for at least 1 week to maximum 6 months) of their participation in a residential recovery program from the 12&12 residential drug addiction treatment center in Tulsa, Oklahoma (Identifier: NCT03382379). Written informed consent was obtained from all participants before participation and the study was approved by the Western IRB (WIRB Protocol #20171742). This study was conducted in accordance with the Declaration of Helsinki and all methods were carried out in accordance with relevant guidelines and regulations. More details on inclusion/exclusion criteria are in supplementary materials (*Section S1*) [42].

### 2.2. Data acquisition procedure

High-resolution structural MRI, resting state fMRI, and fMRI drug cue reactivity task data were collected from all 60 participants on a GE MRI 750 3T scanner. Structural MRI data included T1 and T2-weighted images collected through magnetization-prepared rapid acquisition with gradient-echo (MPRAGE) sequence and were used to create individualized computational head models. During resting-state data collection participants were instructed to relax and stare at a fixation cross, remain awake, and try not to think of anything in particular for 8 minutes. After resting-state data collection, to measure cue-induced brain activity, a standard pictorial block-designed fMRI drug cue reactivity (FDCR) task was administered. Participants were exposed to methamphetamine versus neutral cues (validated in a previous study [43]). The total task time was approximately 6.5 minutes and contained 4 neutral and 4 methamphetamine picture blocks. Each block included a series of 6 pictures of the same category (methamphetamine or neutral), and each was presented for 5 sec with a 0.2-sec blank inter-stimulus interval. A visual fixation point was presented for 8 to 12 sec between each block. More details on MRI data parameters can be found in supplementary materials (*Section S2*). Self-report craving scores were also collected immediately before and after the MRI session using a visual analog scale (VAS) to measure the severity of the craving (0-100).

### 2.3. Task-based functional activity analysis for defining a subcortical seed

Functional data analysis was performed in AFNI. The first three pre-steady state images were removed. The preprocessing steps were as follows: despiking, slice timing correction, realignment, transformation to MNI space, and 4 mm of Gaussian FWHM smoothing. Three polynomial terms and the six motion parameters were regressed-out. TRs with excessive motion (defined as the Euclidian norm of the derivative of the six motion parameters being greater than 0.3) were censored during regression. Statistical analysis of individual imaging data was performed using a first-level fixed effects analysis, in the context of the General Linear Model (GLM). For each participant, a contrast image of methamphetamine > neutral was constructed and a voxel-wise whole-brain analysis was used to identify the effect of the condition (methamphetamine>neutral) and reported P < 0.001. For each participant, Brainnetome atlas parcellation, with a total of 246 cortical and subcortical areas, was applied, and mean beta weight values were estimated for all subregions [44]. The amygdala sub-regions (including left and right medial and lateral amygdala) obtained from the Brainnetime atlas-based parcellation of the fMRI data with the maximum functional activation related to methamphetamine cue reactivity was selected as the seed region that would be modulated by TMS through cortico-subcortical circuits.

### 2.4. Task-based functional connectivity analysis for defining an individualized cortical target

Task-based functional connectivity analysis was performed using the CONN toolbox (v.20.b) [45] in SPM 12. FDCR data underwent a standard preprocessing pipeline in CONN which includes slice timing correction, realignment, co-registration, spatial normalization, and smoothing with an 8mm FWHM Gaussian kernel. Seed-to-whole brain generalized psychophysiological interaction (PPI) analysis (a bivariate regression analysis for task-based connectivity calculations [46]) was performed on the left medial amygdala as the seed. PPI analysis identifies brain regions whose connectivity with the seed region varies as a function of psychological context (here, reactivity to methamphetamine vs. neutral cues). At the first level, the physiological regressors of interest were the timings of methamphetamine and neutral blocks in the task convolved with a hemodynamic response function. The physiological regressor was calculated as the mean time series of the seed region, namely, the left medial amygdala (Brainnetome mask). The PPI regressors were the interaction terms between psychological and physiological regressors, namely, (methamphetamine×physiological) and (neutral×physiological), and the contrast of interest (methamphetamine×physiological) vs (neutral×physiological). These were estimated for each participant to determine which voxels work together with the seed region during the methamphetamine vs. neutral condition. The second level PPI analysis was also performed to determine the significant clusters using voxel-wise and cluster-extent thresholds. Active clusters were reported when surviving a voxel-level statistical threshold of two-sided t-value>3.1 and cluster-level threshold of cluster size>60 voxels.

Based on first-level PPI connectivity maps, we respectively computed personalized TMS targets within the frontopolar area; its mask was obtained from the results reported by Bludau et al. [47]. As there was no specific laterality in previous TMS studies in the field of SUDs, both left and right hemispheres and lateral and medial parts of the frontopolar were searched to find the maximum connection between the amygdala and frontopolar cortex. The voxel with the strongest positive PPI connectivity (Beta values for the PPI regressor as defined in the previous paragraph) to the subcortical area with the highest functional activity during drug cue exposure was selected as the individualized TMS target for each individual.

### 2.5. Construction of head models

Computational head models were generated for all participants using finite element modeling (FEM) implemented in SimNIBS 3.2 standard pipeline [48]. Briefly, T1 and T2 weighted MRI data were used to generate anatomically accurate models based on “headreco” option with the SPM 12 toolbox for tissue segmentation. The constructed head meshes consisted of six tissue types: white matter (WM), gray matter (GM), cerebrospinal fluid (CSF), eyeballs, skull, and scalp with fixed conductivity values; as per the SimNIBS defaults (WM = 0.126 Siemens/meter (S/m), GM = 0.275 S/m, CSF = 1.654 S/m, skull = 0.01 S/m, scalp = 0.465 S/m, and eyeballs = 0.5 S/m) [49]. The results were visualized using Gmsh and MATLAB.

To investigate possible effects due to individualized fMRI-informed TMS targeting, three sets of EF distribution patterns were simulated. (a) F<u>ixed Fp2 approach</u>: in the first set of simulations, EEG 10-10 system Fp2 electrode was selected to target the right frontopolar with fixed standard orientation (y direction toward AF8) across the population in a one-size-fits-all manner (inspired by [27], [50]), (b) <u>Fixed Fp1 approach:</u> in the second set of simulations, EEG 10-10 system Fp1 electrode was selected to target the left frontopolar with fixed standard orientation (y direction towards AF7) across the population in a one-size-fits-all manner (inspired by [16], [18], [19]). These two approaches were commonly used for targeting the frontopolar area in previous TMS studies for SUDs [16], [18], [18], [24], [51]. (c) <u>Optimized approach</u>: optimization was performed at three levels: (1) location, (2) orientation, and (3) intensity. The individualized fMRI-informed approach was used in the third set of simulations. TMS coil was placed over the individualized cortical coordinate obtained from maximum task-based functional connectivity between the amygdala and frontopolar cortex. Coil orientation was then optimized for each person to maximize the EF perpendicular to the targeted brain coordinate using the Auxiliary Dipole Method (ADM) [52] embedded in SimNIBS. These three sets of simulations enabled us to make more direct comparisons between optimized fMRI-informed and fixed TMS targeting, minimizing extraneous methodological differences. Location and value of the 99^th^ percentile of the EFs were calculated for each person and, with respect to the linearity and superposition principle, individualized stimulation intensity was suggested to have similar EF strength (e.g., 1 V/m) in targeted brain regions across the population.

Of the various coil models available in SimNIBS, primary EFs were calculated using a Magstim 70mm Figure-of-eight coil, which is the most common coil type in both clinical and research settings. Mean, median, and peak EF strength were extracted in a 5mm sphere around the individualized targets in each set of simulations. Results were compared using ANOVA with paired sample t-tests as post hoc analyses.

### 2.6. Behavioral analysis

Cue-induced craving was assessed by measuring changes from before to after cue presentation. Self-report craving (“How much craving do you have right now?”) scores were quantified using a visual analog scale (VAS; ranges from 0 to 100, where 0 = no craving and 100 = the highest craving). The correlation between VAS scores and individualized amygdala-frontopolar PPI connectivity strength was calculated using the Pearson correlation coefficient. The study procedure is summarized in *Figure* 1.

**Figure 1.**
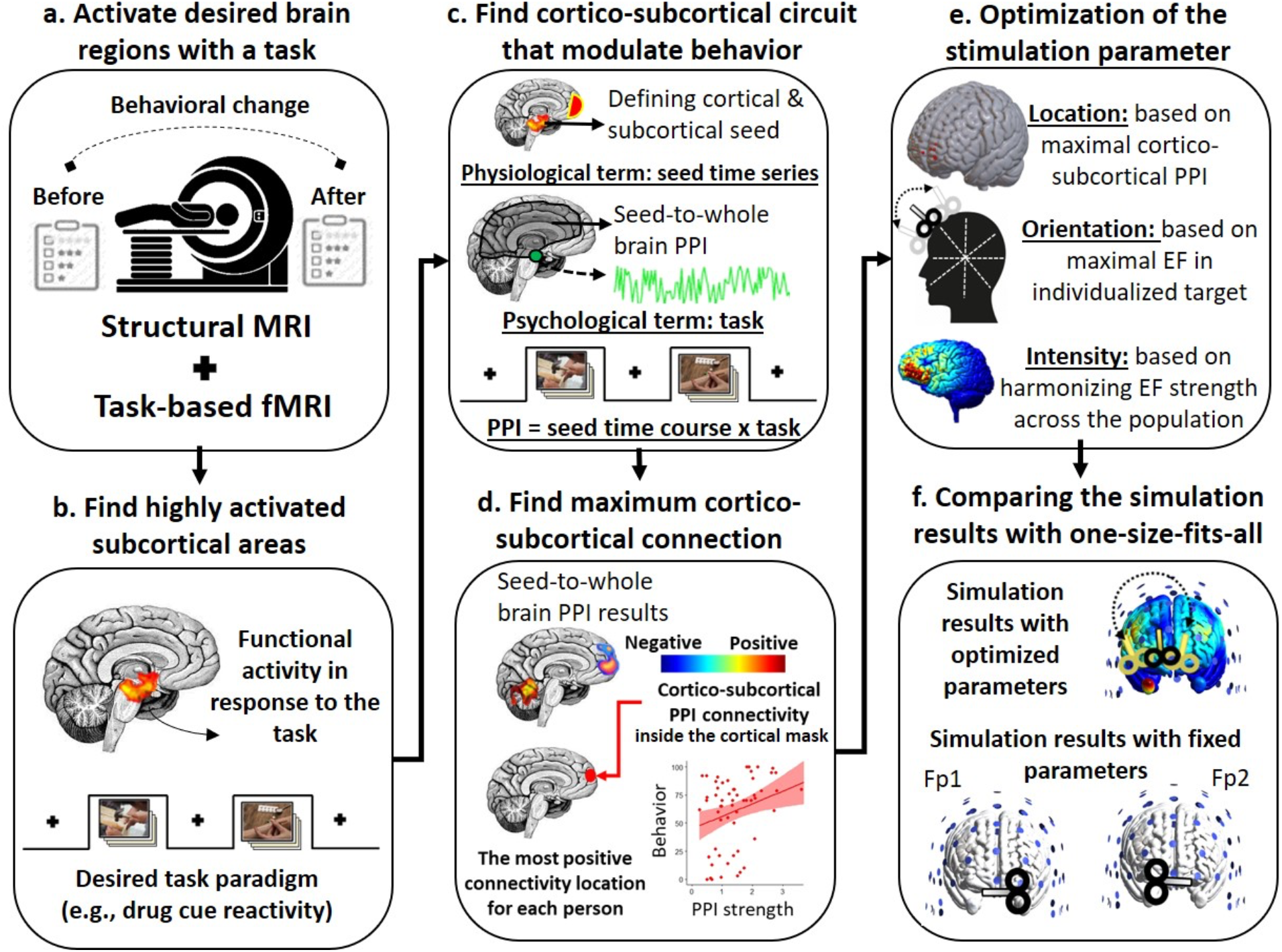
Study procedure. In this study, we used 6 main steps to optimize the TMS protocol for a group of participants with methamphetamine use disorder. **(a) Activating desired brain region with the task of interest**. A standard drug cue reactivity task was used to activate brain regions related to cue-induced craving. Behavioral data including self-report cue-induced craving was also collected immediately before and after the task. **(b) Finding activated subcortical areas**. Functional activity analysis using a general-linear model (GLM) was recruited to find subcortical seed regions that were activated in response to drug vs. neutral cues. **(c) Finding cortico-subcortical circuit that modulates behavior**. Based on a prior hypothesis, the subcortical region with maximal functional activation was selected as the seed. Seed-to-whole brain psychophysiological interaction (PPI) analysis was applied to understand which brain areas are functionally connected to the seed region during drug cue exposure. The cortical target area was selected based on the available literature in the field (e.g., based on the importance of frontopolar-amygdala [53], and previous potential TMS targets [51] in SUD) **(d) Finding max/min cortico-subcortical connection**. The location and strength of the maximal (the most positive value) PPI connection between the subcortical seed region and cortical target mask was extracted for each person. The correlation between maximal PPI strength and changes in cue-induced craving was also calculated. The MNI coordinates of the maximal PPI connections were transformed to the individual space which was considered as the individualized TMS target location for each person. **(e) Optimization of the stimulation parameter**. Computational head models were generated for all participants. The location and orientation of the TMS were optimized for each person based on the maximal PPI locations and maximizing electric field perpendicular to the targeted location. **(f) Comparing the simulation results with one-size-fits-all approaches**. Simulation results obtained from optimized coil location and orientation for each person were compared with two one-size-fits-all approaches that were commonly used for targeting the frontopolar area by placing the TMS coil over the Fp1 or Fp2 locations. Finally, in order to have identical EF strength in the stimulation target, stimulation intensity was adjusted across the population.

## 3. Results

### 3.1. Task-based functional activity results

Functional activity analysis using GLM and Brainnetome atlas parcellation are visualized in *Figure 2*. The level of brain activation (mean ± SD) in methamphetamine>neutral contrast was extracted from each subregion across the sample. Among all subcortical areas (amygdala, hippocamp, thalamus, and basal ganglia), our results based on beta values showed a high level of activation in the amygdala with the highest activity in the left medial amygdala (mean ± SD: 0.31±0.29) ranging from -0.47 to 1.14. The mask of this brain region was selected as a seed for further functional connectivity analysis.

**Figure 2.**
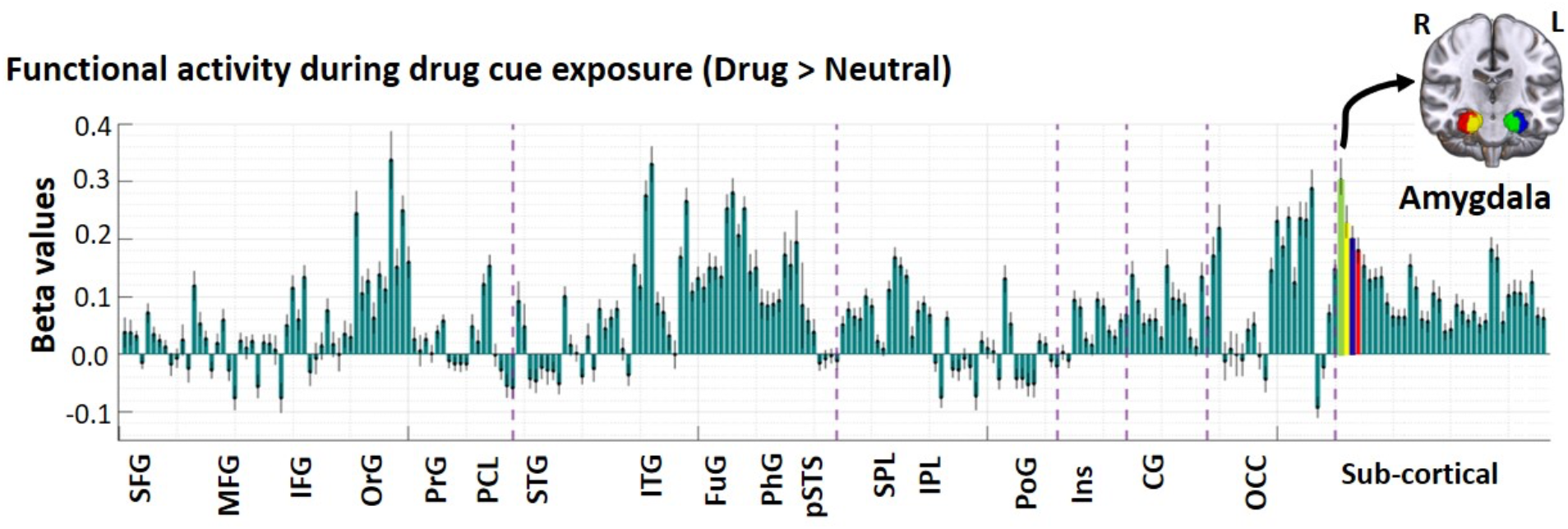
Brain activation during task-based fMRI in Brainnetome atlas parcellation. Using a whole-brain analysis, changes in brain activation during an fMRI drug cue reactivity (FDCR) task in terms of beta values obtained from general linear modeling (GLM) are represented for 60 participants with methamphetamine use disorders. Bars show the mean value and error bars show the standard error of the beta values across the population. As shown, amygdala sub-regions had highest activation among the subcortical areas during cue exposure. Green bar with the highest activation represents the left medial amygdala, yellow bar represents the right medial amygdala, blue bar represents the left lateral amygdala, and red bar represents the right lateral amygdala. Respectively, brain areas over the MNI template represent amygdala subregions based on Brainnetome atlas parcellation. Abbreviation: SFG: superior frontal gyrus, MFG: middle frontal gyrus, IFG: inferior frontal gyrus, OrG: orbital gyrus, PrG: precentral gyrus, PCL: paracentral lobule, STG: superior temporal gyrus, MTG: middle temporal gyrus, ITG: inferior temporal gyrus, FuG: fusiform gyrus, PhG: parahippocampal gyrus, pSTS: posterior superior temporal sulcus, SPL: superior parietal lobule, IPL: inferior parietal lobule, Pcun: precuneus, PoG: postcentral gyrus, Ins: insula, CG: cingulate, OCC: occipital cortex.

### 3.2. Task-based functional connectivity results

Based on the results obtained from functional activity the left medial amygdala was selected as the subcortical seed region. Seed-to-whole brain PPI analyses showed a significant cluster in the right frontopolar cortex survived the voxel-level statistical threshold of two-sided t-value>3.1 and cluster-level threshold of cluster size>60 voxels that showed increased PPI connectivity to amygdala while participants were exposed to drug vs. neutral cues; cluster size: 2462 voxels, cluster peak in MNI space: [25 39 35] (*Figure 3.a*).

**Figure 3.**
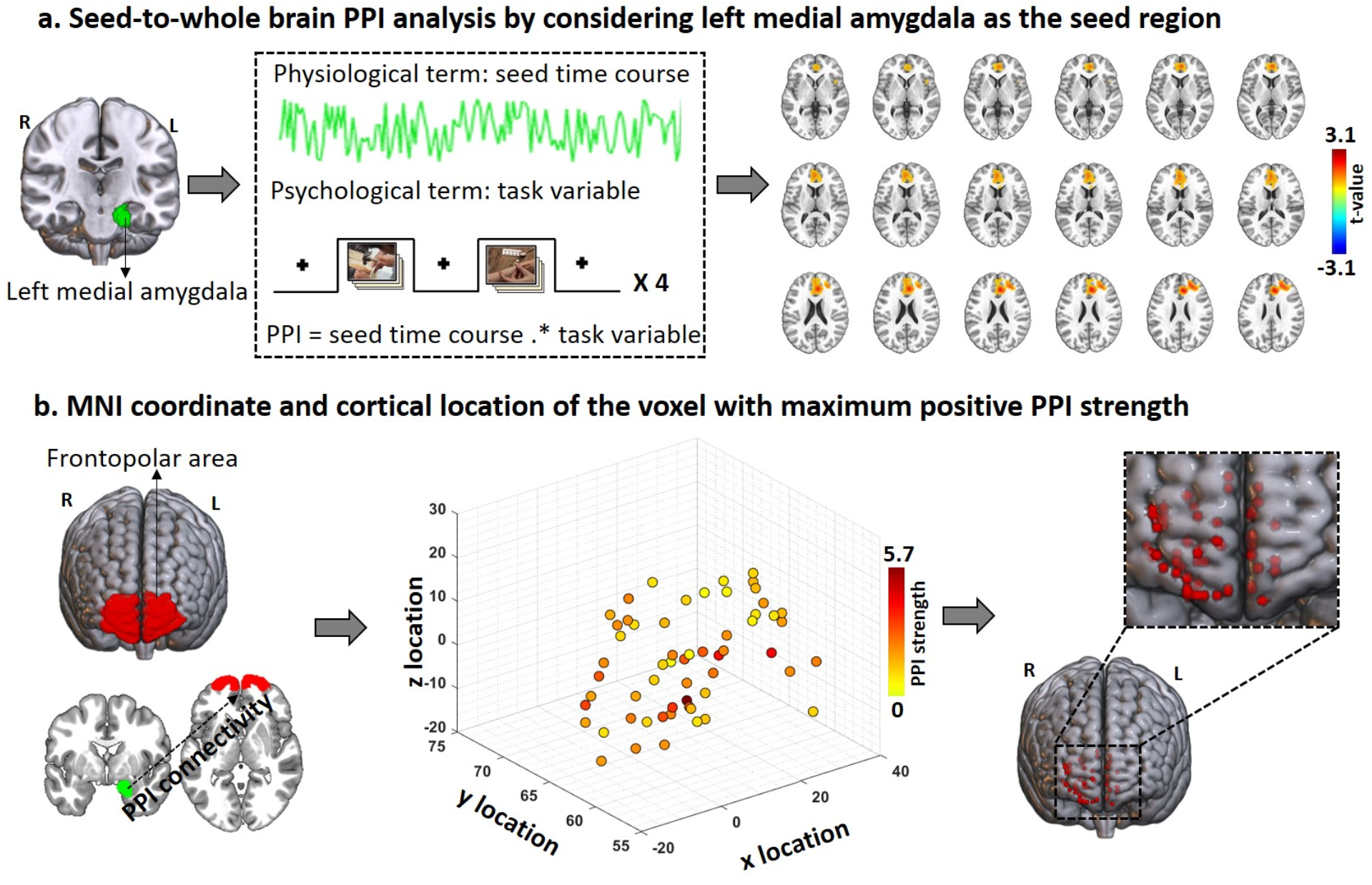
Individualized TMS targets based on amygdala-frontopolar PPI connections. Individualized TMS targets were defined based on psychophysiological interaction (PPI). **(a) Seed-to-whole brain PPI analysis**. The left medial amygdala was selected as the seed region based on the results obtained from functional activity (*Figure 2*). The seed mask extracted from Brainnetome atlas parcellation is visualized in green. Seed-to-whole brain PPI results showed a significant cluster with increased PPI connectivity to the amygdala in the right frontopolar area (cluster size: 2462 voxels, cluster peak in MNI space: [25 39 35]); considering a cluster-level threshold of cluster size>60 voxels. Brain slices in the axial view are shown for better visualization of the significant cluster. **(b) MNI coordinate and cortical location of the voxel with maximum positive PPI strength in the amygdala-frontopolar circuit**. Seed (left medial amygdala in green) and target (frontopolar in red) masks are visualized over the cortical region. Based on the first-level results obtained from the seed-to-whole brain PPI connectivity, the coordinates of the voxel with maximum positive PPI connections inside the right frontopolar mask (the region represented in red) were extracted for each person. In order to visualize inter-individual variability in both location and strength of amygdala-frontopolar connections, scatter plot (for location in MNI space [x y z]) colored based on amygdala-frontopolar PPI strength (beta values in the PPI regression model); hot colors represent strong PPI connectivity strength. The location of the voxel with maximal amygdala-frontopolar PPI connection is visualized over the standard MNI space. Dots represent the data for individual participants. <u>Abbreviation:</u> PPI: psychophysiological interaction.

The coordinate of the maximal PPI connectivity with the left medial amygdala located inside the right medial frontopolar cortex was also determined for each person. Inter-individual variabilities were found in both locations (mean ± SD of the MNI coordinates: [12.60 64.23 -0.8] ± [13.64 3.49 11.01]) and strength (mean ± SD: 1.69 ± 0.96) of the maximal PPI connections across the population. Individualized coordinates were located in a cube with a volume of 63.13mm^3^ and PPI strength (beta values in the bivariate regression defined in section 2.4.) ranged from 0.26 to 5.62 across 60 participants (*Figure 3.b*).

### 3.3. Electric field simulations

Three sets of computational head models were generated for all 60 participants (See section 2.5.). One-way ANOVA showed significant effects for the coil placement approach for all indices (mean (*F* = 50.25, *p* = 2e-16), peak (*F =* 18.08, *p* = 7.7e-8), and median (*F* = 31.77, *p* = 1.98e-12)) extracted from the 5mm spheres. Post hoc pairwise t-test analyses showed that mean EFs in the optimized approach (mean ± SD: 0.99 ± 0.21) were significantly higher than both fixed Fp1 and Fp2 approaches with large effect sizes (Fp1: mean = 0.56 ± 0.22, *p* = 1.1e-13, Hedges’g = 1.50 with 95% of CI (1.12,1.88); Fp2: mean = 0.78 ± 0.25, *p* = 1.7e-5, Hedges’g = 1.26 with 95% of CI (0.91,1.16)). We also found that peak EFs (99^th^ percentiles) in the optimized approach (max = 2.06 ± 0.79) were significantly higher than fixed Fp1 (max = 1.21 ± 0.71, *p* = 5.9e-9) and Fp2 (mean ± SD = 0.78 ± 0.25, *p* = 8.3e-4) with large effect sizes (Fp1: Hedges’g = 1.31 with 95% of CI (0.95,1.66), Fp2: Hedges’g = 1.08 with 95% of CI (0.75,1.41)). Median EF strength was also significantly higher in optimized (median = 0.59 ± 0.17) compared to fixed Fp1 (median = 0.34 ± 0.16, *p* = 8e-5) and Fp2 (median = 0.46 ± 0.18, *p* = 6.2e-5)) with large effect sizes (Fp1: Hedges’g = 1.42 with 95% of CI (1.05,1.79), Fp2: Hedges’g = 1.06 with 95% of CI (0.73,1.38)). Averaged EF strength was also extracted from a 5mm sphere around the individualized targets for each person (ranged from 0.42 to 1.43 V/m). Stimulation intensity was adjusted to have 1 V/m in a 5mm sphere around individualized targeted brain regions across the population.

### 3.4. Behavioral results

Craving scores showed a significant (*p* = 0.00017) increase from before (mean ± SD: 40.95 ± 30.89) to after (61.73 ± 31.11) drug cue exposure with a large effect size (Hedges’g = 0.82 with 95% of CI (0.61,1.03)) (*Figure 5.b*). VAS score after drug cue exposure showed a significant correlation with maximal amygdala-frontopolar PPI connectivity (*R* = 0.27, *p* = 0.034) (*Figure 5.c*). We excluded one participant as an outlier (with PPI = 5.62, VAS-before = 86, VAS-after = 95), however, the Pearson correlation results remained nearly significant (*R* = 0.24, *p* = 0.06).

## Discussion

Here, we explained how structural and functional MRI can be used to identify stimulation parameters for TMS targets and optimize electric field/dose (EF) distribution patterns across a population with methamphetamine use disorder (MUDs) (*Figure 1*). Specifically, this investigation yielded five main results. First, the left medial amygdala showed the highest functional activity during drug cue exposure and was selected as the subcortical seed region. Second, amygdala-to-whole brain PPI analysis showed a significant cluster in the frontopolar/medial prefrontal cortex that confirms cortico-subcortical connections during drug cue exposure. Third, the location of the voxel with the most positive amygdala-frontopolar PPI connectivity in each participant was considered as the individualized TMS target and individual amygdala-frontopolar PPI connectivity in each participant showed a significant correlation with VAS scores after cue exposure. Fourth, the average EF strength in a sphere with r = 5mm around the individualized target location was significantly higher in the optimized compared to the fixed approach with large effect sizes. Finally, stimulation intensity was adjusted for each person to have harmonized to have identical EF intensity over personalized targets for all participants.

### Neural substrate of drug cue reactivity: hyperactivity in the amygdala

Our functional activity analysis of the drug cue reactivity task (*Figure 2*) showed strong activation in all amygdala subregions, with the highest activation in the left medial amygdala while participants were exposed to drug vs. neutral cues. Our results are in accordance with previous studies that suggest the presentation of drug cues appears to reliably activate brain regions involved in the reward/motivation network, including the ventral striatum and amygdala [54], [55], [56]. In this study, the left medial amygdala was chosen as the subcortical seed region due to its highest activation during drug cue exposure. This selection is supported by previous research indicating that amygdala activation in response to drug cue exposure plays a critical role in drug-related behaviors [57]–[59], such that cue-induced craving score is correlated with activation in the amygdala when people were exposed to drug cues [60]. Furthermore, the effectiveness of interventions such as cue exposure therapy [61] or methadone maintenance treatment [62] in reducing cue reactivity in the amygdala provides further support for our seed selection.

### Connections between the amygdala and prefrontal areas in SUDs

Here, we focused on frontopolar-amygdala circuits as a potential TMS target in SUDs and tried to find individualized coordinates to optimally target this brain circuit. Although TMS cannot directly modulate the activity of deeper brain areas like the amygdala, its effects can spread through brain networks [63], [64]. Our amygdala-to-whole brain PPI analysis also supported this idea by revealing a significant positive cluster (increased PPI connections) in the frontopolar/medial prefrontal cortex with a peak activation in the frontopolar cortex (*Figure 3*). This finding confirms cortico-subcortical pathways between the prefrontal amygdala and is in line with previous studies that reported functional connections between the prefrontal cortex and amygdala during drug cue processing [65].

Furthermore, previous studies support our assumption for considering the frontopolar-amygdala circuit as a TMS target for SUDs from different perspectives. (1) Accumulating data highlights the frontopolar cortex as a promising therapeutic target for TMS in SUDs (e.g., lesion-based studies [66], tES studies [67], TMS studies [51]). (2) It has been reported that impairment in functional coupling between the prefrontal cortex and amygdala is negatively correlated with the ability to control negative emotions [68]. (3) Optogenetic studies in male rats showed that chronic alcohol exposure decreased frontopolar-amygdala synaptic strength and accommodation and introduced this brain circuitry as a significant target of the alcohol-associated plasticity [53]. (4) Resting-state fMRI analysis also supported frontopolar-amygdala coupling as a marker of relapse in participants with cocaine use disorders [4]. It has been reported that early relapse risk is associated with reduced connectivity between the left cortico-medial amygdala and the ventromedial prefrontal cortex (comparable to the brain region identified as the frontopolar in our study). In that study, even the site of the amygdala (left medial) is consistent with our results [4]. (5) It has been reported that the frontopolar cortex triggered plasticity events inside the amygdala that plays an important role in the extinction of drug-seeking behaviors [69]. Taken together, all of these findings support the potential for engagement of the amygdala through frontopolar-amygdala functional pathways in the SUD treatment [4], [40], [70], and highlight the necessity of effective TMS protocols for future clinical trials in the field.

### Individualized circuit-based targeting

Coli localization is an important factor in TMS studies. Here, we investigated how far the optimal fMRI-informed target deviates from the fixed EEG-based target which is commonly used. Our individualized approach calculated coordinates for targeting drug-cue-relevant frontopolar-amygdala for each person. In contrast to fixed targeting methods, our results showed notable inter-individual variability in terms of the target locations. Figure 4.a1 showed that this distance was different between individuals. In line with previous findings in connectivity-based target selection [34], [39], these findings reveal variations in the frontopolar coordinate connected to the amygdala, which could potentially explain some of the inter-individual variability observed previously in response to TMS targeting frontopolar cortex for SUDs [71]. Results highlight the importance of coil location optimization in circuit-based stimulation and indicate that anatomical and functional information could be used to estimate the optimal coil position for targeting the frontopolar cortex.

**Figure 4.**
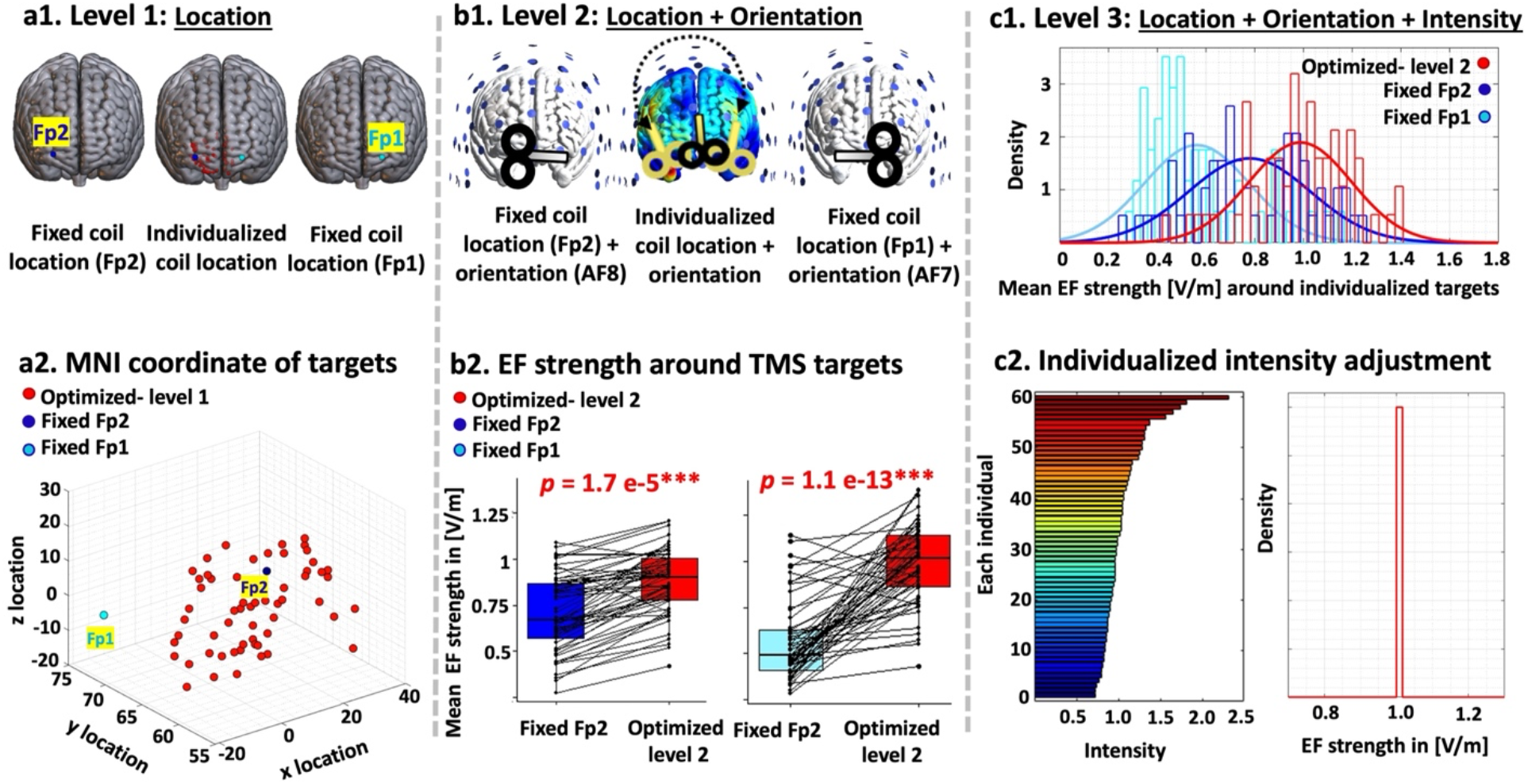
Stimulation parameter optimization. Simulation results were compared between three approaches (1/2) fixed Fp2: coil placed over Fp2/Fp1 in EEG standard system with a fixed orientation toward AF8/AF7 and constant stimulation intensity across the population, and (3) optimized: Optimization was performed at three levels: location, orientation, and intensity. coil was placed over the individualized maximal PPI location, coil direction was optimized to maximize EF perpendicular to the individualized targets, and stimulation intensity was adjusted to have identical 1 V/m averaged EF strength across the population in a 5mm sphere around the individualized targets. **(a) Level 1: Location (coil location individualization)**. Brains in MNI space and the scatter plot shows target location in each approach. The blue dot represents the Fp1 (light blue) and Fp2 (dark blue) locations which were targeted in the fixed approaches and the red dots represent the individualized targets in the optimized approach obtained from maximal amygdala-frontopolar PPI connectivity. **(b) Level 2: Location + Orientation (coil orientation optimization in individualized targets)**. In the fixed approaches standard coil directions were used (AF8/AF7) while in the optimized approach optimization algorithm (ADM method) tried to maximize perpendicular EFs in individualized brain targets. Boxplots showing effects of orientation optimization on averaged EF in a 5mm sphere around the individualized target. Dots and spaghetti lines over the boxplots represent the data for each individual participant. P values for differences are reported above boxplots. **(c) Level 3: Location + Orientation + Intensity (stimulation intensity adjustment)**. Distributions of the averaged EF strength in a 5mm sphere around the individualized targets are represented for the fixed (Fp1/Fp2 in light/dark blue) and optimized (in red) approaches. With respect to linearity and superposition principles, EF distribution patterns with optimized level2 parameters were simulated after correction of the stimulation intensity to have 1 V/m in a 5mm sphere around individualized targeted brain regions across the population. The adjustment factor for each individual is represented in the lower part of panel c (hot color represents more strong adjustment factors). <u>Color code:</u> light blue: Fp1, dark blue: Fp2, red: Optimized. <u>Abbreviation:</u> EF: electric field, PPI: psychophysiological interaction, V/m: volt per meter.

We considered the strongest positive z-value in the connection between the amygdala and frontopolar as the TMS target. Because it has been reported that inhibitory rTMS over the frontopolar can potentially revert hyperactivity in brain subcortical regions such as the amygdala [19], [26], [28], and based on the common frequency-dependent effects of rTMS, we expect that inhibitory stimulation to a positively connected frontopolar node would dampen the positive connectivity between the frontopolar and amygdala, a change that could potentially reduce drug-related behaviors like drug craving and consumption. We also found a positive significant correlation between the most positive amygdala-frontopolar PPI connectivity and self-reported craving scores after drug cue exposure (*Figure* 5, people with stronger connectivity reported a higher cue-induced craving score). This finding supports our assumption and suggests that inhibitory stimulation over the coordinates with the most positive connections to the amygdala might be successful in reducing craving scores. Future research should include clinical testing of different TMS targeting methods to understand the comparative effects of each technique on behavioral outcomes.

**Figure 5.**
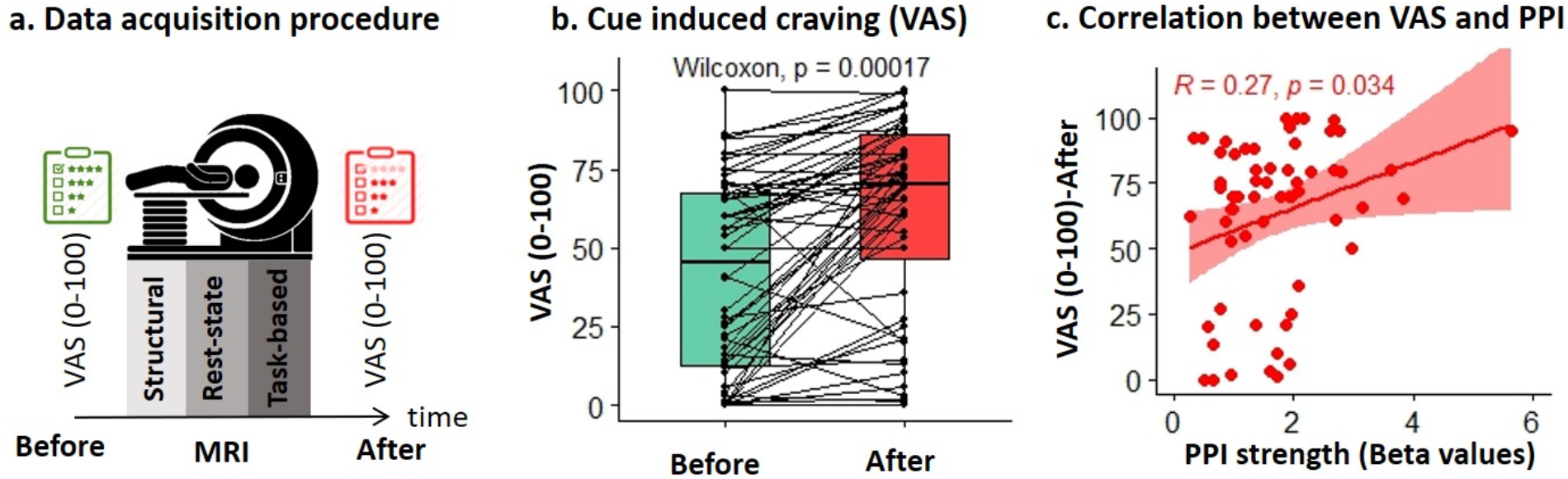
Cue-induced craving changes and amygdala-frontopolar PPI connectivity. (a) Data acquisition procedure. Self-report craving scores based on visual analog scale (VAS) were collected immediately before (in green) and after (in red) MRI session (started with T1 and T2-weighted structural MRI and followed by resting-state fMRI which was followed by fMRI drug cue reactivity task data collection). **(b) Cue-induced craving**. Boxplots showing effects of time (before and after MRI session) on drug craving. Dots and spaghetti lines over the boxplots represent the data for each individual participant. P values for differences between VAS before and after scanning are reported above boxplots. **(c) VAS-PPI Correlation**. The association between VAS scores after cue exposure and maximal amygdala-frontopolar PPI connectivity showed a significant correlation. Dots represent each individual and the boundary is an indicator of the 95% confidence interval. R and P values are reported for the Pearson correlation coefficient between VAS scores after the MRI session and maximal PPI strength between the left medial amygdala and the frontopolar cortex. <u>Color code:</u> green: VAS scores before cue exposure, red: VAS scores after cue exposure, <u>Abbreviation:</u> VAS: visual analog scale, PPI: psychophysiological interaction.

### Optimizing coil orientation

Here, we optimized coil orientation over personalized TMS targets in the frontopolar area. Due to the presence of measurable markers such as motor-evoked potential, the optimal coil orientation is well-known for the motor cortex while the optimal coil orientation remains to be determined for other brain areas [72]. Furthermore, although there is an increasing number of fMRI-informed TMS studies to determine coil location for each person, less attention has been paid to optimizing coil orientation in previously published clinical trials [36], [38], [73]. Our results showed that coil orientation optimization significantly increased EF strength over the frontopolar cortex compared to fixed orientation, which is commonly used in TMS studies. Our results suggest that coil angle optimization based on the anatomical shape of the head should be considered for estimating the most effective stimulation parameters over the frontopolar cortex as supported by previous TMS studies in other brain targets like motor cortex [72], [74].

### Harmonizing EF strength across the sample

Our simulations with a fixed TMS dose showed inter-individual variability at the cortical target site (*Figure 4*). In line with previous modeling studies, at the cortical target site, in both fixed and individualized fMRI-informed locations, intensities substantially varied between individuals [75], [76]. Our findings support that fixed-dose TMS which is typically used in clinical trials may result in varying physiological or behavioral effects among individuals. As suggested by Evans et al controlling for this source of variability must be a priority [77]. Therefore, we adjusted the stimulation intensity for each individual to control the stimulation dose and reduce EF variability in the targeted brain region. As expected, by individualizing the dose, the EF was consistent near the target site and reached a specific target intensity. Here, inspired by [77], we adjusted stimulation intensity to harmonize EF across individuals with the assumption that there is a straightforward (linear) dose-response relationship between the physiological/behavioral effects of TMS and the delivered stimulation dose in the targeted brain region. Hence, we controlled the EF that was delivered to each personalized target location. Our assumption is supported by previous studies suggesting that EF intensity in a cortical target predicts response to the applied brain stimulation technology [78], [79]. However, other factors such as EF direction in the targeted area, the spatial extent of the EF in each person, above threshold EF intensity in non-targeted brain regions, or ongoing brain state may be relevant to the observed response.

### Limitations and future directions

Our study has some limitations that could be addressed in future research. First, whether frontopolar-amygdala connectivity is the clinically optimal target for MUDs remains an open question. Although there are promising results in previous frontopolar-amygdala connectivity-based TMS in healthy young adults [40], the fMRI-informed target selection approach is still new in the field of brain stimulation for SUDs [80]. Replication in larger cohorts to draw a general conclusion about the optimal cortico-subcortical biomarker is needed that can be informative for predictive/treatment-response biomarker extraction in future TMS studies.

Second, our study did not include manipulation of the targeted circuit with TMS. Prospective research testing the proposed optimized targeting approach needs to be tested in future studies by collecting fMRI before and after optimized rTMS and comparing the results with a non-optimized approach (e.g., fixed standard method) to test for BOLD and connectivity changes due to stimulation and how each approach affects behavioral outcomes, e.g., craving.

Third, we only investigated task-based fMRI data to engage the fronto-limbic network and induce Hebbian-like plasticity [81]. Others have focused on resting-state fMRI data collection [82], [83] to reduce the need for participant compliance and avoid confounds related to task performance or instructions [84]. fMRI-informed TMS target selection based on resting-state networks may exhibit higher reproducibility compared to conventional task-based imaging [85]. Direct comparisons of task-based vs. resting-state-defined TMS network targeting will be important in future SUD research.

Fourth, we created computational head models only based on T1 and T2-weighted images with previously established isotropic conductivities, as is common in computational studies. Anisotropic skull and white matter conductivities significantly affect EF distribution patterns [86] which is important when considering deeper target regions inside the brain [87]. Anisotropic conductivities could be calculated based on diffusion tensor imaging in future studies to increase the accuracy of computational head modeling as it has been shown that in TMS simulations anisotropy led to differences up to 10% in the maximum induced EFs [88]. Furthermore, we used constant tissue conductivities while variations in reported human head tissue electrical conductivity values may have effects on simulation outcomes [89]. Although assuming conductivity from the literature is insufficient, obtaining the personalized values at body temperature at frequencies<100Hz and in natural, in-vivo conditions would be difficult and unavailable. Other physiological factors such as motor-evoked potentials that could be partly explained by EFs in the motor cortex can be used for future dose-controlled TMS studies [79].

We focused only on coil location, orientation, and stimulation intensity harmonization across the population. However, there is a large TMS parameter space, and many other factors can affect stimulation outcomes that could be considered in future studies. For example, in a TMS clinical trial, train duration, inter-train interval, pulse numbers, session number, pulse width, pulse shape, and frequency could be optimized for each individual by considering initial brain state, EF distribution patterns, and behavioral outcomes.

Here, our sample only consisted of male participants. Previous studies on brain stimulation and substance use disorders have reported on the impact of sex differences on electric field distribution patterns and brain functions [90], [91]. It is recommended to include both males and females in future research to ensure the findings are representative of the entire population.

Future dose-response studies could be informative to optimize all related factors for each individual or across a population based on the association between the stimulation parameter and stimulation efficacy [92], [93]. For example, an inverted U-shaped dose-response curve was reported by Huang et al., for the number of stimulation pulses such that 600 pulses produced a more durable response than 300, while 1200 pulses produced inhibitory effects rather than increasing the effects of 600 pulses [94]. Dose-response relationship studies can be leveraged to design new TMS parameters that optimally manipulate brain responses in future clinical trials.

Finally, future closed-loop TMS-fMRI systems that allow automatic adjustment of the stimulation location and orientation, e.g., with a robotically controlled stimulator, raise the possibility of real-time optimization of the coil location, orientation, and stimulation intensity/frequency based on personalized ongoing brain state. Automated closed-loop TMS-fMRI with a suitable optimization algorithm implementation help to adaptively adjust stimulation parameters in a multidimensional search space [95]. However, an MRI-compatible device, a high-speed optimization algorithm, and powerful computational resources for generating and analysis of head models are needed to implement closed-loop TMS-fMRI systems.

## Conclusion

The effectiveness of non-invasive brain stimulation technologies like TMS is limited by individual differences in response. This study aimed to improve TMS outcomes by customizing stimulation parameters based on individual variability of functional connectivity parameters. The study found that personalized TMS targets can improve connectivity between the amygdala and frontopolar circuits, which are important for regulating addiction-related behaviors. Furthermore, our proposed circuit was consistent with a previously reported circuit that was identified as a predictor of relapse status [4]. Thus, the study emphasizes the importance of individualized coil placement, orientation, and stimulation intensity adjustment. These findings can help refine TMS therapy for substance use disorders, and future research should investigate whether this approach can improve neural and behavioral outcomes in clinical trials.

## Data Availability

All data produced in the present study are available upon reasonable request to the authors.

## Acknowledgement

Authors declared no conflict of interest.

## Funding

This study is supported by funds from Laureate Institute for Brain Research, Tulsa, OK, Medical Discovery Team on Addiction, University of Minnesota, Minneapolis, MN and Brain and Behavior Foundation (NARSAD Young Investigator Award #27305) to HE. The grant RF1MH117428 was provided to AO. There was no role for the funding agency in the design, execution, analysis or reporting this study.

## Supplementary materials

### S1. Inclusion/Exclusion criteria

The inclusion criteria for this study were: (1) English speaking, (2) diagnosed with methamphetamine use disorder in the last 12 months, (3) admitted to a residential abstinence-based treatment program for methamphetamine use disorder, (4) abstinence from methamphetamine for at least one week, and (5) willing and capable of interacting with the informed consent process. Exclusion criteria included: (1) unwillingness or inability to complete any of the major aspects of the study protocol, including magnetic resonance imaging (i.e. due to claustrophobia), drug cue rating or behavioral assessment, (2) abstinence from methamphetamine for more than 6 months based on self-report, (3) schizophrenia or bipolar disorder based on the MINI interview, (4) active suicidal ideation with intent or plan determined by self-report or assessment by the principal investigator or study staff during the initial screening or any other phase of the study, (5) positive drug test for amphetamines, opioids, cannabis, alcohol, phencyclidine, or cocaine confirmed by breath analyzer and urine tests.

### S2. MRI data parameters

<u>Structural MRI parameters:</u> TR/TE = 5/2.012 ms, FOV/slice = 24 × 192/0.9 mm, 256×256 matrix producing 0.938 × 0.9 mm voxels and 186 axial slices for T1-weighted images and TR/TE=8108/137.728ms, FOV/slice=240/2mm, 512×512 matrix producing 0.469×0.469×2mm voxels and 80 coronal slices for T2-weighted images. T1- and T2-weighted MR images were used for generating computational head models (CHMs) for each individual. <u>Resting-state fMRI parameters:</u> TR/TE = 2000/27 ms, FOV/slice = 240/2.9 mm, 128×128 matrix producing 1.857×1.857×2.9 mm voxels, 39 axial slices, and 240 repetitions. <u>Task-based fMRI parameters:</u> had the same parameters in resting-state, except containing 196 repetitions.

## Notes

### Competing Interest Statement

The authors have declared no competing interest.

### Clinical Trial

NCT03382379

### Author Declarations

Written informed consent was obtained from all participants before participation and the study was approved by the Western IRB (WIRB Protocol #20171742).

### Summary of Updates

three new references are added and the funding section is updated.

